# A proposed general variant classification framework using chronic pancreatitis as a disease model

**DOI:** 10.1101/2022.06.03.22275950

**Authors:** Emmanuelle Masson, Wen-Bin Zou, Emmanuelle Génin, David N. Cooper, Gerald Le Gac, Yann Fichou, Na Pu, Vinciane Rebours, Claude Férec, Zhuan Liao, Jian-Min Chen

**Author notes:** These authors share co-first authorship (Emmanuelle Masson, Wen-Bin Zou). **Correspondence:** Jian-Min Chen, MD, PhD, INSERM UMR1078 – EFS – UBO, 22 avenue Camille Desmoulins, 29238 BREST, France.

## Abstract

The widely used ACMG-AMP variant classification categories (pathogenic, likely pathogenic, uncertain significance, likely benign and benign) were specifically developed for variants in Mendelian disease genes, classifying variants discretely with respect to a simple causal versus benign dichotomy. A general variant classification framework taking into account the continuum of clinical phenotypes, the continuum of the variants’ genetic effects and the different pathological roles of the genes implicated, is however lacking. Herein, we used chronic pancreatitis (CP), which clinically manifests as hereditary, familial, idiopathic or alcoholic forms, as a disease model. Based upon cross-gene and cross-variant comparisons, we firstly assigned the four most studied CP genes (*PRSS1, CFTR, SPINK1* and *CTRC*) to two distinct categories in terms of causality: CP-causing (*PRSS1* and *SPINK1*) and CP-predisposing (*CFTR* and *CTRC*). We then employed two new classificatory categories, “predisposing” and “likely predisposing”, to replace ACMG-AMP’s “pathogenic” and “likely pathogenic” categories in CP-predisposing genes, thereby classifying all pathologically relevant variants in these genes as “predisposing”. In the case of CP-causing genes, the two new classificatory categories served to expand the five ACMG-AMP categories whilst two thresholds (allele frequency and functional) were introduced to discriminate pathogenic from predisposing variants. Our proposed five-category (predisposing, likely predisposing, uncertain significance, likely benign and benign) and seven-category (pathogenic, likely pathogenic, predisposing, likely predisposing, uncertain significance, likely benign and benign) frameworks (with respect to disease-predisposing and disease-causing genes, respectively) retain the backbone of the five ACMG-AMP categories while rendering them readily applicable to variant classification in other disease contexts.

## Introduction

With the routine application of exome and genome sequencing in clinical settings, we are facing an increasing challenge in terms of assigning variants to the five discrete classificatory categories (i.e., “pathogenic”, “likely pathogenic”, “uncertain significance”, “likely benign” and “benign”)^1^ recommended by the American College of Medical Genetics and Genomics and the Association for Molecular Pathology (ACMG-AMP). The key issue is that the ACMG-AMP guidelines were specifically drawn up in order to describe variants identified in genes underlying Mendelian disorders. However, in reality, the etiology of a given disorder may (i) lie on a spectrum from highly penetrant single gene defect to multifactorial disease and (ii) involve multiple gene loci that do not have the same pathological impact on the disease in question. Moreover, even in genes underlying Mendelian disorders, clinically relevant variants do not readily fall into a discontinuous causal versus benign dichotomy.^2^ Indeed, as opined by Wright and colleagues,^3^ some basic conceptual questions about variant interpretation still remain to be addressed in medical genetics. Thus, should the term “pathogenic” be generally applied to any disease-relevant variant in a given disease-causing gene? When should a pathologically relevant mutation be considered to be a “risk” variant rather than being “pathogenic” in its own right? Various adaptations and refinements of the ACMG-AMP guidelines have previously been made in the context of secondary findings derived from clinical exome and genome sequencing^4^ as well as in different gene/disease or variant contexts^5-11^ but none provides a general framework that addresses the aforementioned conceptual issues.

Chronic pancreatitis (CP), a chronic inflammatory process of the pancreas that leads to irreversible morphological changes and the progressive impairment of both exocrine and endocrine functions, can be caused by both genetic and environmental factors.^12,13^ In common with many other diseases, genetic discoveries in CP started with the mapping and identification of a causative gene for a Mendelian form of the disease, autosomal dominant hereditary pancreatitis.^14^ Thereafter, a diverse range of variants in more than 10 different genes (for references, see Masson et al.^15^) have been identified in patients with hereditary, familial, idiopathic and/or alcoholic CP (see Methods for disease subtype definitions) that may be considered to reflect a continuum of the disease extending from monogenic to multifactorial.^16^ Herein, using CP as a disease model (Figure 1), we have attempted to provide a general variant classification framework, by adapting and extending ACMG-AMP guidelines.

**Figure 1.**
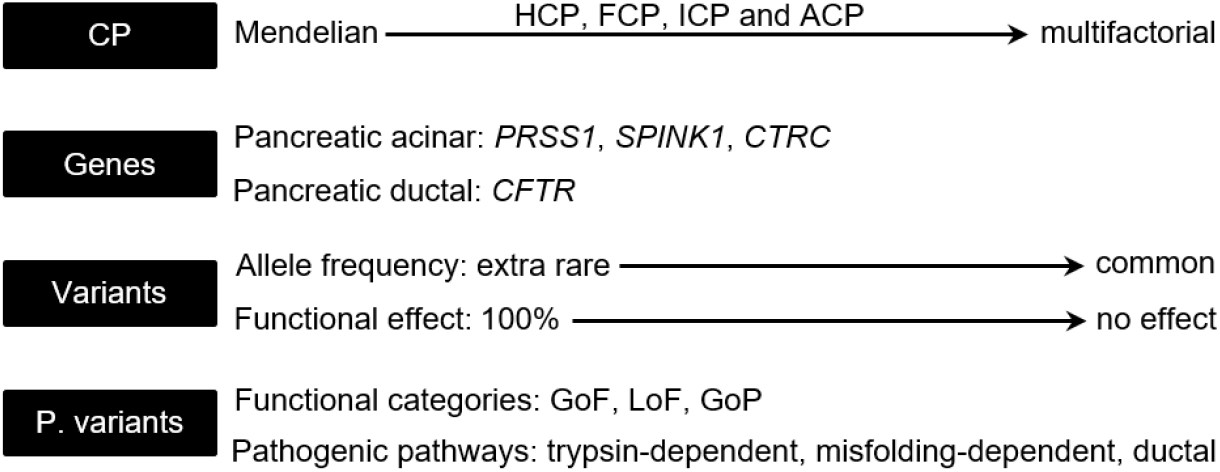
Layers of complexity challenging variant classification in CP that were included for analysis in the current study. CP, chronic pancreatitis; HCP, hereditary CP; FCP, familial CP; ICP, idiopathic CP; ACP, alcoholic CP; P. variants, pathological variants; GoF, gain-of-function; LoF, loss-of-function; GoP, gain-of-proteotoxicity.

## Results

### Genes included for analysis

A general conceptual proposal should be based upon well-established knowledge. We therefore opted to focus our analysis on the first discovered and most extensively studied four CP genes, *PRSS1, CFTR, SPINK1* and *CTRC*, each harboring a large number of pathologically relevant variants.^14,17-27^ General information about these four genes, including the encoded protein, year and approach of gene discovery, mRNA reference accession number, length of coding DNA sequence and length of the encoded protein, may be found in Table 1.

**Table 1.**
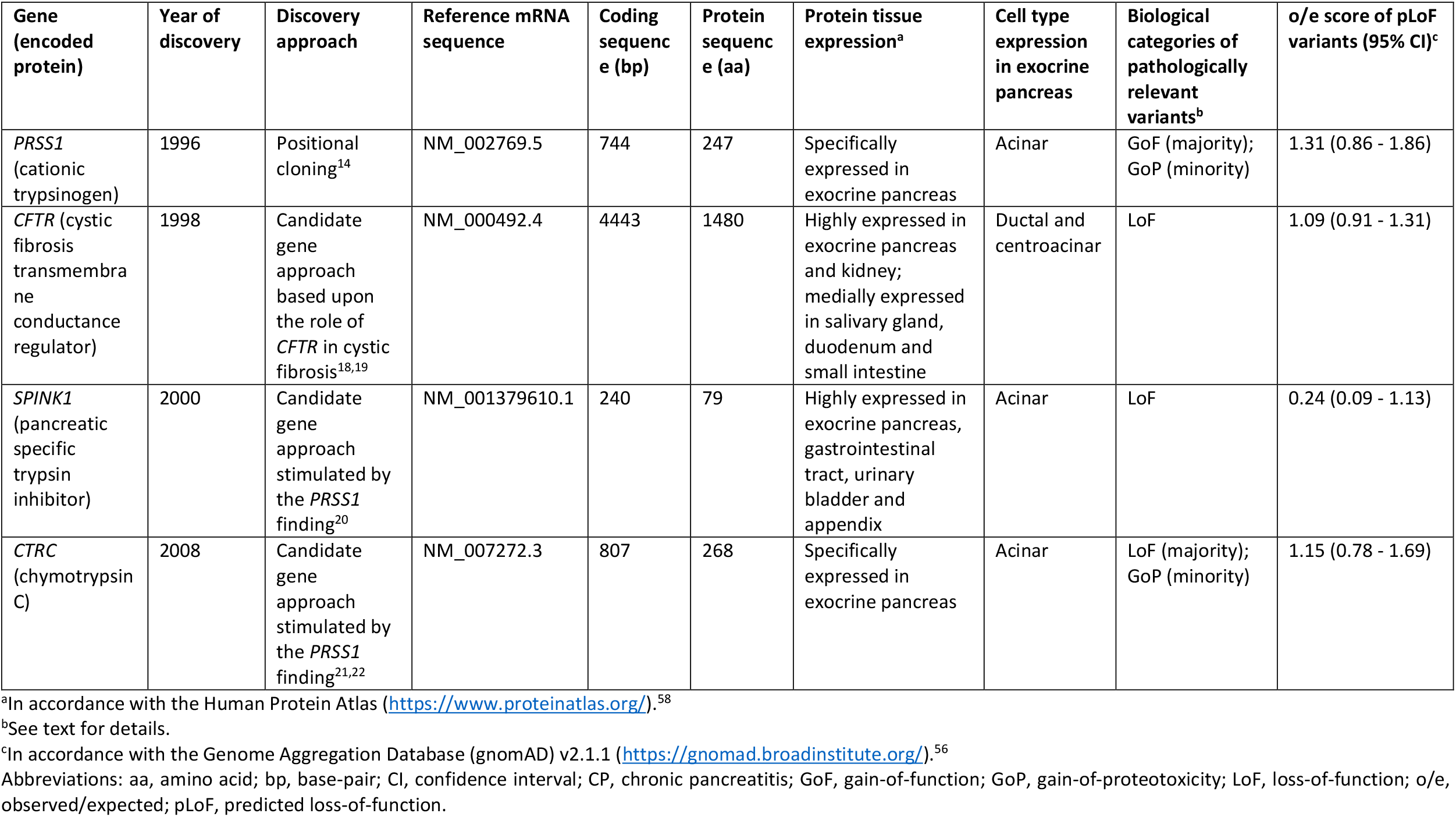
Some general information about the four CP genes.

| Gene (encoded protein) | Year of discovery | Discovery approach | Reference mRNA sequence | Coding sequence (bp) | Protein sequence (aa) | Protein tissue expression<sup>a</sup> | Cell type expression in exocrine pancreas | Biological categories of pathologically relevant variants<sup>b</sup> | o/e score of pLoF variants (95% CI)<sup>c</sup> |
| --- | --- | --- | --- | --- | --- | --- | --- | --- | --- |
| *PRSS1* (cationic trypsinogen) | 1996 | Positional cloning<sup>14</sup> | NM_002769.5 | 744 | 247 | Specifically expressed in exocrine pancreas | Acinar | GoF (majority); GoP (minority) | 1.31 (0.86 - 1.86) |
| *CFTR* (cystic fibrosis transmembrane conductance regulator) | 1998 | Candidate gene approach based upon the role of *CFTR* in cystic fibrosis<sup>18,19</sup> | NM_000492.4 | 4443 | 1480 | Highly expressed in exocrine pancreas and kidney; medially expressed in salivary gland, duodenum and small intestine | Ductal and centroacinar | LoF | 1.09 (0.91 - 1.31) |
| *SPINK1* (pancreatic specific trypsin inhibitor) | 2000 | Candidate gene approach stimulated by the *PRSS1* finding<sup>20</sup> | NM_001379610.1 | 240 | 79 | Highly expressed in exocrine pancreas, gastrointestinal tract, urinary bladder and appendix | Acinar | LoF | 0.24 (0.09 - 1.13) |
| *CTRC* (chymotrypsin C) | 2008 | Candidate gene approach stimulated by the *PRSS1* finding<sup>21,22</sup> | NM_007272.3 | 807 | 268 | Specifically expressed in exocrine pancreas | Acinar | LoF (majority); GoP (minority) | 1.15 (0.78 - 1.69) |

<sup>a</sup>In accordance with the Human Protein Atlas (https://www.proteinatlas.org/).<sup>58</sup>
<sup>b</sup>See text for details.
<sup>c</sup>In accordance with the Genome Aggregation Database (gnomAD) v2.1.1 (https://gnomad.broadinstitute.org/).<sup>56</sup>
Abbreviations: aa, amino acid; bp, base-pair; CI, confidence interval; CP, chronic pancreatitis; GoF, gain-of-function; GoP, gain-of-proteotoxicity; LoF, loss-of-function; o/e, observed/expected; pLoF, predicted loss-of-function.

### Classifying the pathologically relevant variants in the four CP genes into three categories in terms of their biological consequences

*PRSS1, SPINK1* and *CTRC* are specifically or highly expressed in the acinar cells whereas *CFTR* is highly expressed in the ductal cells of the exocrine pancreas (Table 1). Based upon current knowledge, all pathologically relevant variants in the four CP genes could be classified into three biological categories: gain-of-function (GoF), loss-of-function (LoF) and gain-of-proteotoxicity (GoP). Briefly, GoF variants in *PRSS1* result in increased trypsinogen activation and/or increased trypsin stability. These variants, as well as LoF variants in *SPINK1* and *CTRC* (NB. SPINK1 specifically inhibits trypsin whereas CTRC specifically degrades trypsinogen/trypsin), give rise to increased intrapancreatic trypsin activity or a gain of *trypsin* within the pancreas, thereby causing or predisposing to CP (trypsin-dependent pathway).^28,29^ A small subset of pathologically relevant variants in *PRSS1* and *CTRC* induced the misfolding of their corresponding zymogens and elicited endoplasmic reticulum (ER) stress in the pancreatic acini (misfolding-dependent pathway);^30^ these variants are termed GoP. In the exocrine pancreas, CFTR regulates cAMP-mediated bicarbonate secretion into the pancreatic duct lumen, which dilutes and alkalinizes the protein-rich acinar secretions; LoF variants in CFTR are thought to lead to slowed flushing of trypsinogen/trypsin out of the pancreatic ducts, thereby predisposing to pancreatic injury and CP^16^ (termed ‘ductal pathway’ by Mayerle and colleagues^31^). These classifications served as the basis to perform the cross-gene and cross-variant comparisons outlined below.

### Classifying the four CP genes into two distinct categories in terms of causality

The four CP genes do not contribute equally to the pathophysiology of the exocrine pancreas. To distinguish their roles in the pathogenesis of CP at the gene level, we firstly determined whether the very rare variants (defined as having a minor allele frequency (MAF) of <0.001 in accordance with Manolio et al.^32^ in any gnomAD subpopulations) were identified in the Mendelian form of CP or hereditary CP (HCP) in the context of each gene. A MAF cutoff of 0.001 has previously been recommended for filtering variants responsible for dominant Mendelian disorders.^33^ The MAF of <0.001 corresponds to a carrier frequency of <0.002. It was used here as a very conservative cutoff given that it was more than 600 times higher than the prevalence of HCP, which was estimated to be 0.3/100 000 in Western Countries.^34^ The premise was that such variants, where presumed (or experimentally demonstrated) to fall into the aforementioned GoF, GoP or LoF categories, can be confidently interpreted as disease-causing.

*PRSS1* was the first CP gene to be identified, with multiple very rare variants including GoF copy number and missense variants and GoP missense variants being reported in many HCP families. By contrast, only a limited number of very rare *SPINK1* variants, and not particularly very rare *CFTR* and *CTRC* variants, have been reported in HCP families. Moreover, the HCP families harboring *PRSS1* mutations were generally large, often involving ≥4 patients across ≥3 generations whereas the HCP families harboring *SPINK1* mutations had at the most 3 patients over 3 generations (Table 2). In short, high-confidence disease-causing variants were found in *PRSS1* and *SPINK1* but not in *CFTR* and *CTRC*.

**Table 2.** Very rare pathologically relevant variants found in HCP in the context of four CP genes.

| Gene | Variant<sup>a</sup> | Number of HCP families (family description) reported<sup>b</sup> | Reference(s) | Biological/functional consequence | gpAF (hspAF) in gnomAD<sup>c</sup> |
| --- | --- | --- | --- | --- | --- |
| *PRSS1* | Trypsinogen gene triplication | 5 (10 patients across 4 generations<sup>d</sup>) | Le Maréchal et al.<sup>17</sup> | GoF (gene dosage)<sup>29</sup> | Absent |
|  | Double “gain-of-function” hybrid variant | 1 (6 patients across 3 generations) | Masson et al.<sup>59</sup> | GoF (gene dosage plus effect of p.Asn29Ile) | Absent |
|  | c.47C>T (p.Ala16Val) | 2 (4 patients across 2 generations; 3 patients across 2 generations) | Grocock et al.<sup>60</sup> | GoF (increased activation)<sup>61</sup> | Absent<sup>57</sup> |
|  | c.62A>C (p.Asp21Ala) | 1 (5 patients across 3 generations) | Yilmaz et al.<sup>62</sup> | GoF (increased activation)<sup>63</sup> | Absent |
|  | c.63_71dup (p.Lys23_Ile24insIleAspLys) | 1 (3 patients across 2 generations) | Joergensen et al.<sup>64</sup> | GoF (increased activation)<sup>64</sup> | Absent |
|  | c.86A>T (p.Asn29Ile) | The second most frequent variant causing HCP;<sup>44</sup> in the first report, one family had 19 patients across 7 generations.<sup>65</sup> | Gorry et al.<sup>65</sup> | GoF (increased activation and stability)<sup>61</sup> | Absent |
|  | c.86A>C (p.Asn29Thr) | 1 (8 patients across 3 generations) | Dytz et al.<sup>66</sup> | GoF (increased activation and stability)<sup>61</sup> | Absent |
|  | c.116T>C (p.Val39Ala) | 1 (9 patients across 3 generations) | Arduino et al.<sup>67</sup> | GoF (increased stability)<sup>61</sup> | Absent |
|  | c.311T>C (p.Leu104Pro) | 2 (both having 3 patients across 3 generations) | Teich et al.<sup>68</sup>; Németh et al.<sup>69</sup> | GoP (intracellular retention and elevation of ER stress marker)<sup>70</sup> | Absent |
|  | c.346C>T (p.Arg116Cys) | 2 (3 patients across 2 generations; 3 patients across 3 generations) | Pho-lam et al.<sup>71</sup>; Kereszturi et al.<sup>72</sup> | GoP (intracellular retention and elevation of ER stress marker)<sup>72</sup> | 0.00007072 (0.0007018, East Asian) |
|  | c.365G>A (p.Arg122His) | The most frequent variant causing HCP;<sup>44</sup> in the discovery report, one family had 20 patients across 4 generations.<sup>14</sup> |  | GoF (increased activity)<sup>73,74</sup> | 0.00001194 (0.00002639, non-Finnish European) |
|  | c.365_366GC>AT<br>(p.Arg122His) | 1 (4 patients across 4 generations) | Howes et al.<sup>75</sup> | Same as above | Absent |
| *CFTR* | Not identified |  |  |  |  |
| *SPINK1* | c.27DelC<br>(p.Ser10ValfsTer5) | 1 (3 patients across 2 generations) | Le Maréchal et al.<sup>76</sup> | LoF (predicted complete functional loss) | 0.00001197<br>(0.00002896,<br>Latino/Admixed<br>American) |
|  | c.41T>G (p.Leu14Arg) | 2 (both having 3 patients across 3 generations) | Király et al.<sup>77</sup> | LoF (experimentally demonstrated to abolish SPINK1 secretion)<sup>77</sup> | Absent |
|  | Deletion of the entire gene | 1 (3 patients across 2 generations) | Masson et al.<sup>78</sup> | LoF (predicted complete functional loss) | Absent |
| *CTRC* | Not identified |  |  |  |  |

<sup>a</sup>All are heterozygous. See Table 1 for reference mRNA accession numbers.
<sup>b</sup>Data from some original reports were reinterpreted in accordance with our working definition of HCP, specified in the Methods section.
<sup>c</sup>In accordance with gnomAD v2.1.1 or SVs v2.1 (https://gnomad.broadinstitute.org/).<sup>56</sup>
<sup>d</sup>Described was the family with the most affected patients.
Abbreviations: CP, chronic pancreatitis; gpAF, global population allele frequency; GoF, gain-of-function; GoP, gain-of-proteotoxicity; HCP, hereditary CP; hspAF, highest subpopulation allele frequency; LoF, loss-of-function.

The abovementioned findings may have been affected by many factors including differences in patient recruitment and mutation analysis protocols between laboratories and different timespans since the first report of CP gene discovery. To confirm or refute these findings, we performed three additional comparative analyses. Firstly, we compared the observed/expected (o/e) scores of predicted LoF (pLoF) variants in the four genes from gnomAD v2.1.1 (Table 1). The o/e score is an indicator of LoF intolerance devised by Karczewski and colleagues,^35^ low o/e values being indicative of strong intolerance. The highest o/e score was exhibited by *PRSS1* (1.31); this is understandable because it is predominantly GoF variants in this gene that cause CP whereas LoF variants in *PRSS1* and *PRSS2* (encoding anionic trypsinogen, the second major isoform after cationic trypsinogen) are protective with respect to CP.^36,37^ With regard to the latter, we evaluated the pLoF *PRSS1* variants in gnomAD v2.1.1. The highest subpopulation allele frequency (hspAF) of such variants, which was found in the case of the c.200+1G>A variant, is 0.02871 (African/African American). Notably, in the context of the three genes for which LoF variants (or predominantly LoF variants) underlie the disease, *CFTR* and *CTRC* have an o/e score of >1 (1.09 and 1.15, respectively) whereas *SPINK1* has an o/e score of <1 (specifically, 0.24).

Secondly, we compared the odds ratios (ORs) calculated from the aggregated pathologically relevant variants in the three genes for which LoF variants (or predominantly LoF variants) underlie the disease. For reasons of simplicity and comparability, we used data from a German study that analyzed these genes in a large cohort of patients (n = 410-660) and controls (n = 750-1758).^24^ The ORs for *CFTR, CTRC* and *SPINK1* variants were 2.7, 5.3 and 15.6, respectively. In other words, the aggregated pathologically relevant variants in the *CFTR* and *CTRC* genes had a much lower genetic effect than those in the *SPINK1* genes.

Thirdly, reinforcing the above point, even the most severe LoF variants in *CFTR* and *CTRC* do not exert a very large genetic effect. Thus, for example, *CFTR* p.Phe508del, the classical cystic fibrosis-causing variant, had an OR of only 2.5 (95% CI 1.7-3.9) for CP.^24^ In similar vein, *CTRC* p.Lys247_Arg254del, which causes a complete loss of enzymatic activity, had an OR of 6.4 (95% CI 2.3-17.5).^38^ By contrast, the OR for idiopathic CP (ICP) conferred by *SPINK1* c.194+2T>C, which should result in a ∼90% functional loss of SPINK1,^39,40^ was 59.31 (95% CI 33.93-103.64) based upon data from a Chinese study.^26,41^

Finally, a remarkable difference in terms of phenotype expression was observed between human *SPINK1* and *CTRC* knockouts. Two *SPINK1* knockouts, one a homozygous deletion of the entire *SPINK1* gene, the other the homozygous insertion of a full-length inverted *Alu* element into the 3’-untranslated region of the *SPINK1* gene (experimentally determined to cause the complete loss of *SPINK1* expression), presented with severe exocrine pancreatic insufficiency around 5 months of age.^42^ By contrast, a *CTRC* knockout, homozygous for a deletion of the entire *CTRC* locus, had been clinically asymptomatic until adulthood.^43^ Only at the age of 20 was he incidentally found to have calcifications and cysts in the pancreas; subsequent laboratory tests revealed exocrine pancreatic insufficiency.^43^ These highly unusual cases are strongly consistent with SPINK1 exerting a much stronger effect than CTRC in terms of the negative regulation of the level of prematurely activated trypsin within the pancreas.

Taking these observations together, we classified *PRSS1* and *SPINK1* as CP-causing genes and *CFTR* and *CTRC* as CP-predisposing genes.

### Adapting ACMG-AMP guidelines for the classification of variants in the two CP-predisposing genes

We would propose simply to change two of the five ACMG categories, “pathogenic” and “likely pathogenic”, to “predisposing” and “likely predisposing” for the purposes of classifying the pathologically relevant variants in the *CFTR* and *CTRC* genes (Figure 2a). Thus, all *CFTR* pathologically relevant variants previously known as “cystic fibrosis-causing, severe”, “cystic fibrosis-causing, mild” and “non cystic fibrosis-causing”^24^ will be classified as “predisposing” in the context of CP, with the conventional cystic fibrosis-based categories being provided in parentheses. As for the *CTRC* variants, all “pathogenic” variants listed in the Genetic Risk Factors in Chronic Pancreatitis Database^44^ will be reclassified as CP “predisposing”.

**Figure 2.**
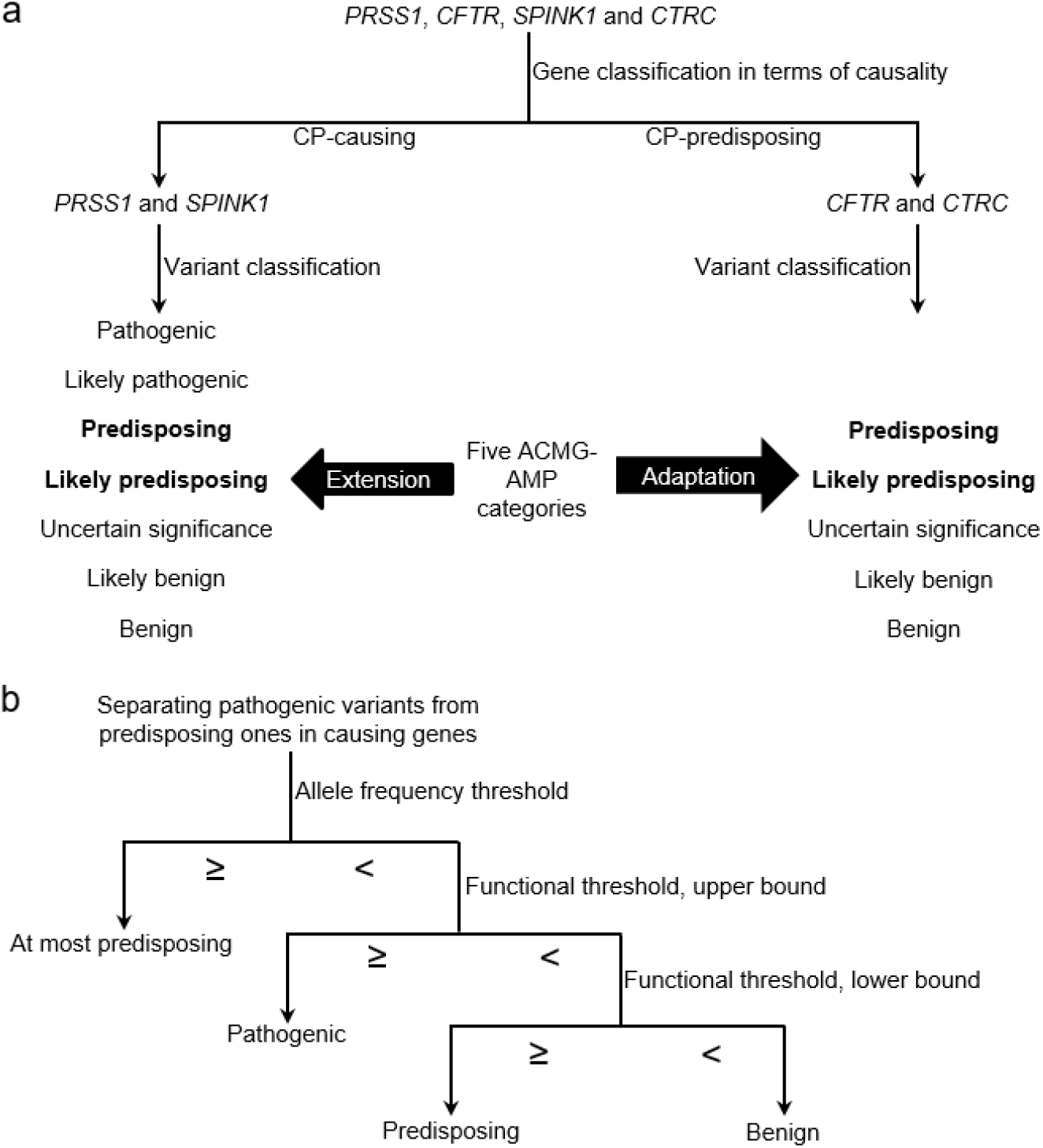
Two key components of our proposed variant classification framework. (**a**) Disease genes were first classified into either ‘causing’ or ‘predisposing’ based upon multiple layers of evidence. Then, minimal extension and adaptation were made to the five ACMG-AMP categories in the different gene contexts. The two new categories proposed in this study are highlighted in bold. (**b**) Illustration of using two thresholds for distinguishing pathogenic variants from predisposing variants in disease-causing genes. CP, chronic pancreatitis

### Extending the ACMG guidelines to classify variants in the two CP-causing genes

Evidently, not all pathologically relevant variants in a disease-causing gene are causative. To make a distinction at this juncture, we propose to add the above-mentioned two novel categories, “predisposing” and “likely predisposing”, to the five ACMG categories (Figure 2a). Therefore, the key issue is how to distinguish “pathogenic” from “predisposing” among the pathologically relevant variants in the causative genes (Figure 2b).

#### Establishing an allele frequency threshold to distinguish pathogenic variants from predisposing ones

The rarity of a variant is a proxy indicator of potential pathogenicity. But defining an allele frequency threshold above which a pathological variant should be considered too common to cause the disease in question is challenging due to uncertainties pertaining to disease prevalence on the one hand and the existence of genetic and allelic heterogeneity as well as the issue of incomplete penetrance on the other.^45^

Earlier, we used a conservative MAF cutoff of <0.001 to evaluate high confidence HCP-causing variants. Herein, we further explore this issue by evaluating the population allele frequencies of what we term “gold-standard” pathologically relevant variants in the two CP-causing genes. “Gold-standard” LoF variants in *SPINK1* refer to pLoF variants or variants experimentally shown to cause a complete or almost complete (>95%) loss of SPINK1 function. By contrast, it is impractical to quantify the effect of GoF or GoP variants. Keeping this *caveat* in mind, “gold-standard” GoF variants in *PRSS1* refer to those variants that are very rare and which have been experimentally shown to increase trypsinogen activation and/or trypsin stability whereas “gold-standard” GoP variants in *PRSS1* refer to those variants that are very rare and which have experimentally been shown to reduce protein secretion and elicit ER stress. The global population allele frequency (gpAF) and hspAF of these “gold-standard” variants are provided in Tables 3-5. Herein, it should be noted that in the context of “gold-standard” LoF variants in *SPINK1* (Table 5), p.Arg67His, which was experimentally shown to cause a complete functional loss of SPINK1,^46^ has a hspAF as high as 0.03078. This apparent outlier was excluded from the final analysis.

**Table 3.**
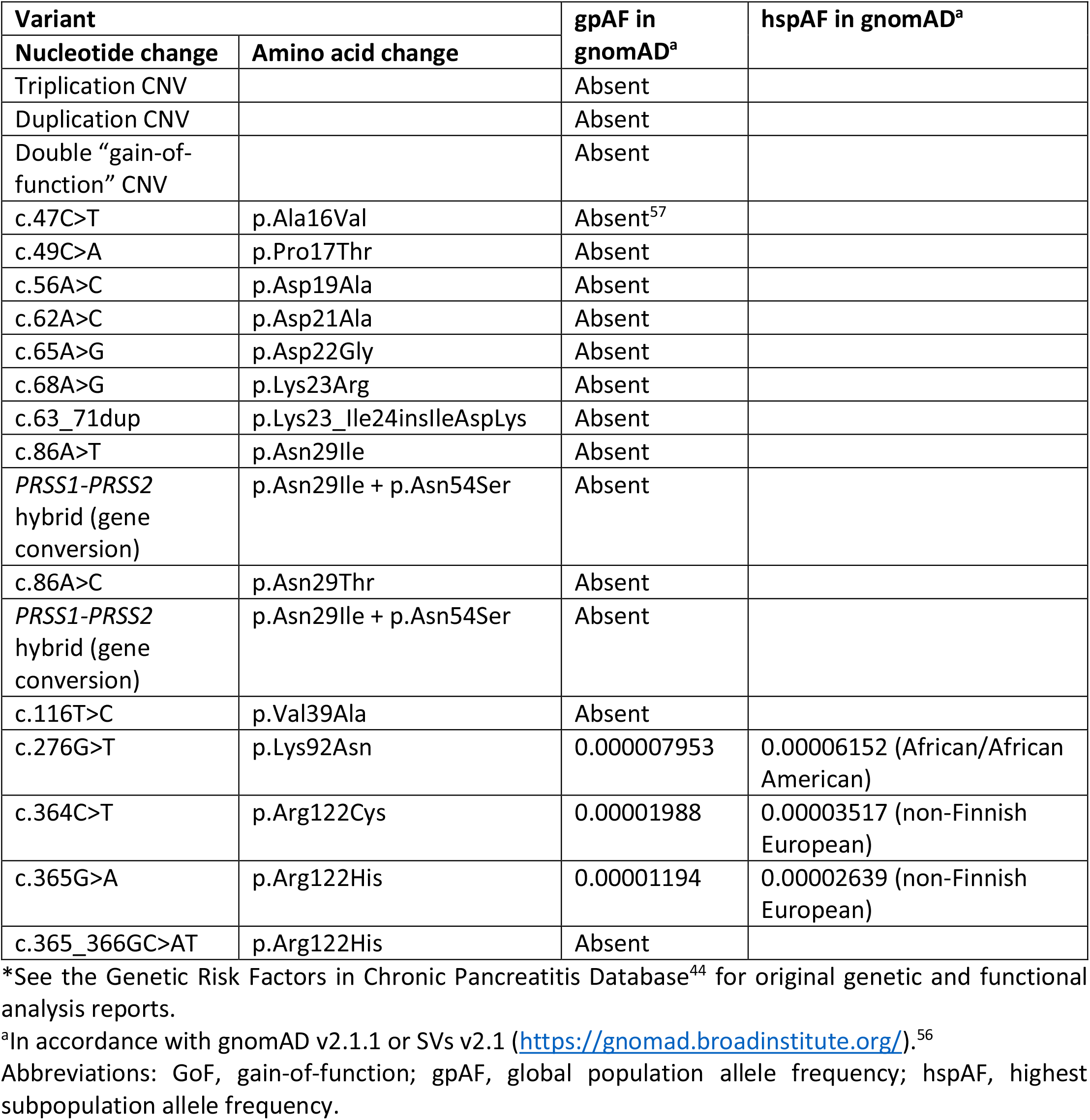
“Gold-standard” GoF variants in *PRSS1\**.

**Table 4.** “Gold-standard” GoP variants in *PRSS1\**.

| Variant |  | gpAF in gnomAD<sup>a</sup> | hspAF in gnomAD<sup>a</sup> |
| --- | --- | --- | --- |
| Nucleotide change | Amino acid change |  |  |
| c.311T>C | p.Leu104Pro | Absent |  |
| c.346C>T | p.Arg116Cys | 0.00007072 | 0.0007018 (East Asian) |
| c.415T>A | p.Cys139Ser | Absent |  |
| c.416G>T | p.Cys139Phe | Absent |  |

\*See the Genetic Risk Factors in Chronic Pancreatitis Database<sup>44</sup> for original genetic and functional analysis reports.
<sup>a</sup>In accordance with gnomAD v2.1.1 or SVs v2.1 (https://gnomad.broadinstitute.org/).<sup>56</sup>
Abbreviations: GoP, gain-of-proteotoxicity; gpAF, global population allele frequency; hspAF, highest subpopulation allele frequency.

**Table 5.**
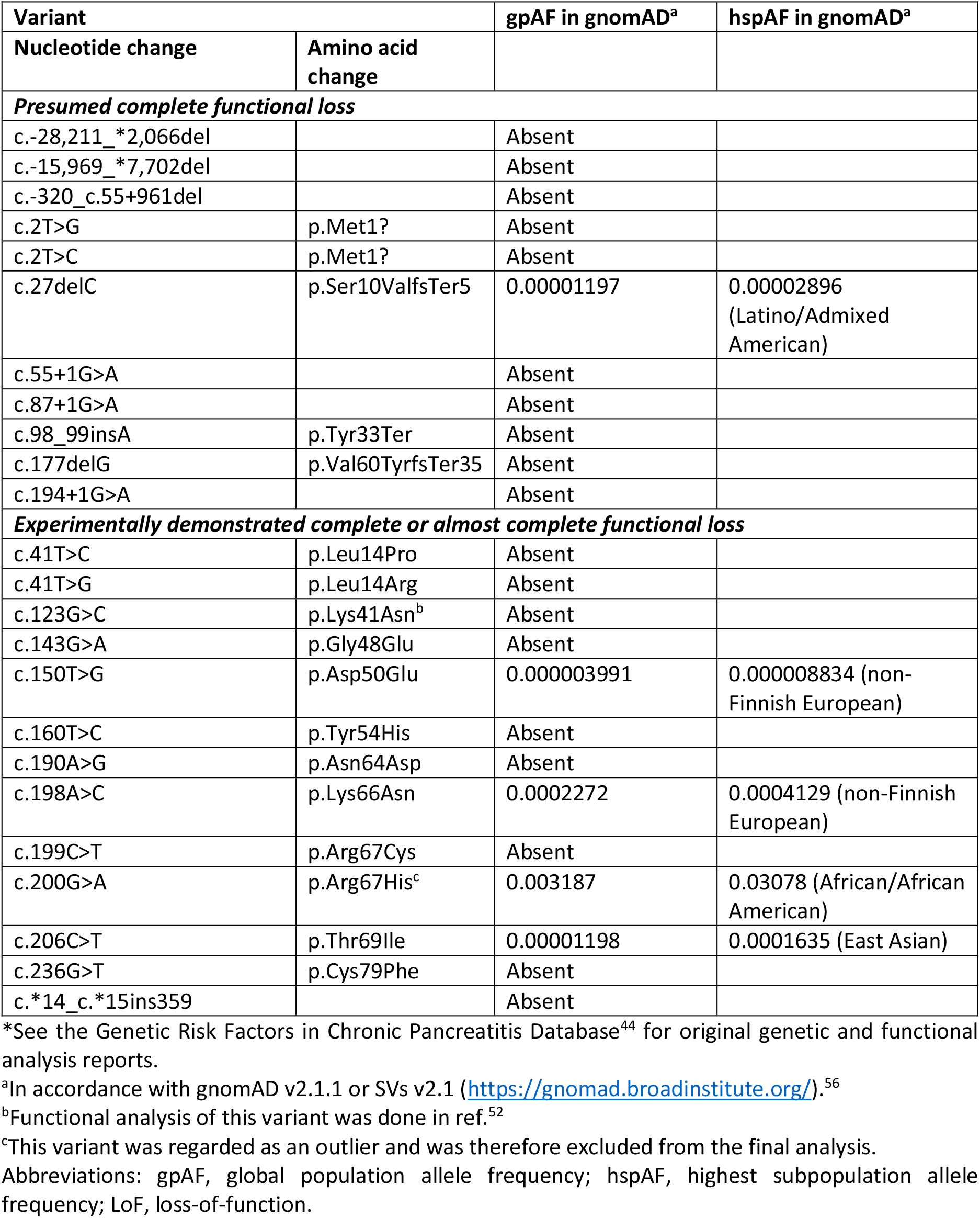
“Gold-standard” LoF variants in *SPINK1\**.

| Variant |  | gpAF in gnomAD<sup>a</sup> | hspAF in gnomAD<sup>a</sup> |
| --- | --- | --- | --- |
| Nucleotide change | Amino acid change |  |  |
| ***Presumed complete functional loss*** |  |  |  |
| c.-28,211_*2,066del |  | Absent |  |
| c.-15,969_*7,702del |  | Absent |  |
| c.-320_c.55+961del |  | Absent |  |
| c.2T>G | p.Met1? | Absent |  |
| c.2T>C | p.Met1? | Absent |  |
| c.27delC | p.Ser10ValfsTer5 | 0.00001197 | 0.00002896<br>(Latino/Admixed American) |
| c.55+1G>A |  | Absent |  |
| c.87+1G>A |  | Absent |  |
| c.98_99insA | p.Tyr33Ter | Absent |  |
| c.177delG | p.Val60TyrfsTer35 | Absent |  |
| c.194+1G>A |  | Absent |  |
| ***Experimentally demonstrated complete or almost complete functional loss*** |  |  |  |
| c.41T>C | p.Leu14Pro | Absent |  |
| c.41T>G | p.Leu14Arg | Absent |  |
| c.123G>C | p.Lys41Asn<sup>b</sup> | Absent |  |
| c.143G>A | p.Gly48Glu | Absent |  |
| c.150T>G | p.Asp50Glu | 0.000003991 | 0.000008834 (non-Finnish European) |
| c.160T>C | p.Tyr54His | Absent |  |
| c.190A>G | p.Asn64Asp | Absent |  |
| c.198A>C | p.Lys66Asn | 0.0002272 | 0.0004129 (non-Finnish European) |
| c.199C>T | p.Arg67Cys | Absent |  |
| c.200G>A | p.Arg67His<sup>c</sup> | 0.003187 | 0.03078 (African/African American) |
| c.206C>T | p.Thr69Ile | 0.00001198 | 0.0001635 (East Asian) |
| c.236G>T | p.Cys79Phe | Absent |  |
| c.*14_c.*15ins359 |  | Absent |  |

\*See the Genetic Risk Factors in Chronic Pancreatitis Database<sup>44</sup> for original genetic and functional analysis reports.
<sup>a</sup>In accordance with gnomAD v2.1.1 or SVs v2.1 (https://gnomad.broadinstitute.org/).<sup>56</sup>
<sup>b</sup>Functional analysis of this variant was done in ref.<sup>52</sup>
<sup>c</sup>This variant was regarded as an outlier and was therefore excluded from the final analysis.
Abbreviations: gpAF, global population allele frequency; hspAF, highest subpopulation allele frequency; LoF, loss-of-function.

As shown in Tables 3-5, only a small subset of “gold-standard” pathologically relevant variants in the two CP-causing genes were found in normal populations. Of this small set of variants, the high-confidence HCP-causing *PRSS1* p.Arg116Cys has the highest hspAF (0.0007018). We elect to adopt the previously recommended allele frequency of 0.001 for the filtering of dominant Mendelian disorders^33^ as the threshold hspAF for differentiating pathogenic from predisposing variants in the *PRSS1* and *SPINK1* genes.

#### Establishing gene-specific functional thresholds to distinguish pathogenic variants from predisposing ones

In the two CP-causing genes, not all pathologically relevant variants with a hspAF of <0.001 can be pathogenic due to their different functional effects. Taking into consideration the different roles of the two genes, we attempted to set gene-specific functional thresholds that would allow pathogenic variants to be distinguished from predisposing ones.

As mentioned earlier, it is impractical to quantify the effect of GoF or GoP variants in the *PRSS1* gene. Given the central role of PRSS1 in the trypsin-dependent pathway and that *PRSS1* is the most abundantly expressed of the pancreatic zymogen genes, we would tentatively classify all *PRSS1* variants, with an allele frequency of <0.001 that were experimentally demonstrated to be consistent with a GoF or GOP mechanism, as pathogenic.

We would further propose that *SPINK1* variants with an allele frequency of <0.001, that were either presumed or experimentally shown to cause a complete or almost complete functional loss (>95%) of SPINK1, should be regarded as pathogenic. Additional support for this proposition came from the *SPINK1* c.194+2T>C variant which is associated with a ∼90% functional loss of SPINK1^39,40^ but has an hspAF of 0.003335 in the East Asian population. As for the lower bound of functional loss for defining predisposing *SPINK1* variants, we would tentatively propose a functional loss of at least 10%.

#### Employing the two newly established thresholds to reclassify several variants in the two CP-causative genes

In the Genetic Risk Factors in Chronic Pancreatitis Database,^44^ variants in the *PRSS1* and *SPINK1* genes were systematically classified in accordance with the ACMG-AMP recommended five categories with the addition of a new “protective” category. Herein, we mainly focus on the missense variants and pLoF variants that were classified as “pathogenic” or “likely pathogenic” in *PRSS1* and *SPINK1* by the Database.^44^ Utilizing the newly established thresholds would result in the reclassification of multiple variants;

In the context of *PRSS1*, p.Gly208Ala would be reclassified from “pathogenic” to “predisposing”, primarily because its hspAF is 0.00987 (East Asian), ∼10 times higher than the 0.001 threshold; moreover, functional assays revealed that this variant had only a moderate impact on secretion;^47^ furthermore, in terms of its genetic effect, it had an OR of only 4.92 for ICP.^26,41^ “Pathogenic” p.Lys92Asn and p.Ser124Ser variants would be reclassified as “likely pathogenic” since both of them showed moderate impact on secretion but no data on ER stress were available. We also propose to reclassify the “protective” LoF variants p.Tyr37Ter and c.200+1G>A as “benign”, with a view to avoiding the addition of a clinically irrelevant category to the five pre-existing ACMG-AMP categories. Nonetheless, to distinguish them from the classical “benign” variants (e.g., missense variants that have been experimentally demonstrated to be functionally neutral), the “protective” nature of these LoF variants in *PRSS1* may be specified in parentheses after the “benign” category (Table 6). Employing the same line of reasoning, we would propose to use the risk allele rather than the protective allele for variant classification with respect to the common promoter variant located at c.-204, upstream of the translational initiation codon of *PRSS1*.^48-50^ Consequently, c.-204C>A (protective) should be described as c.-204A>C (predisposing).

**Table 6.**
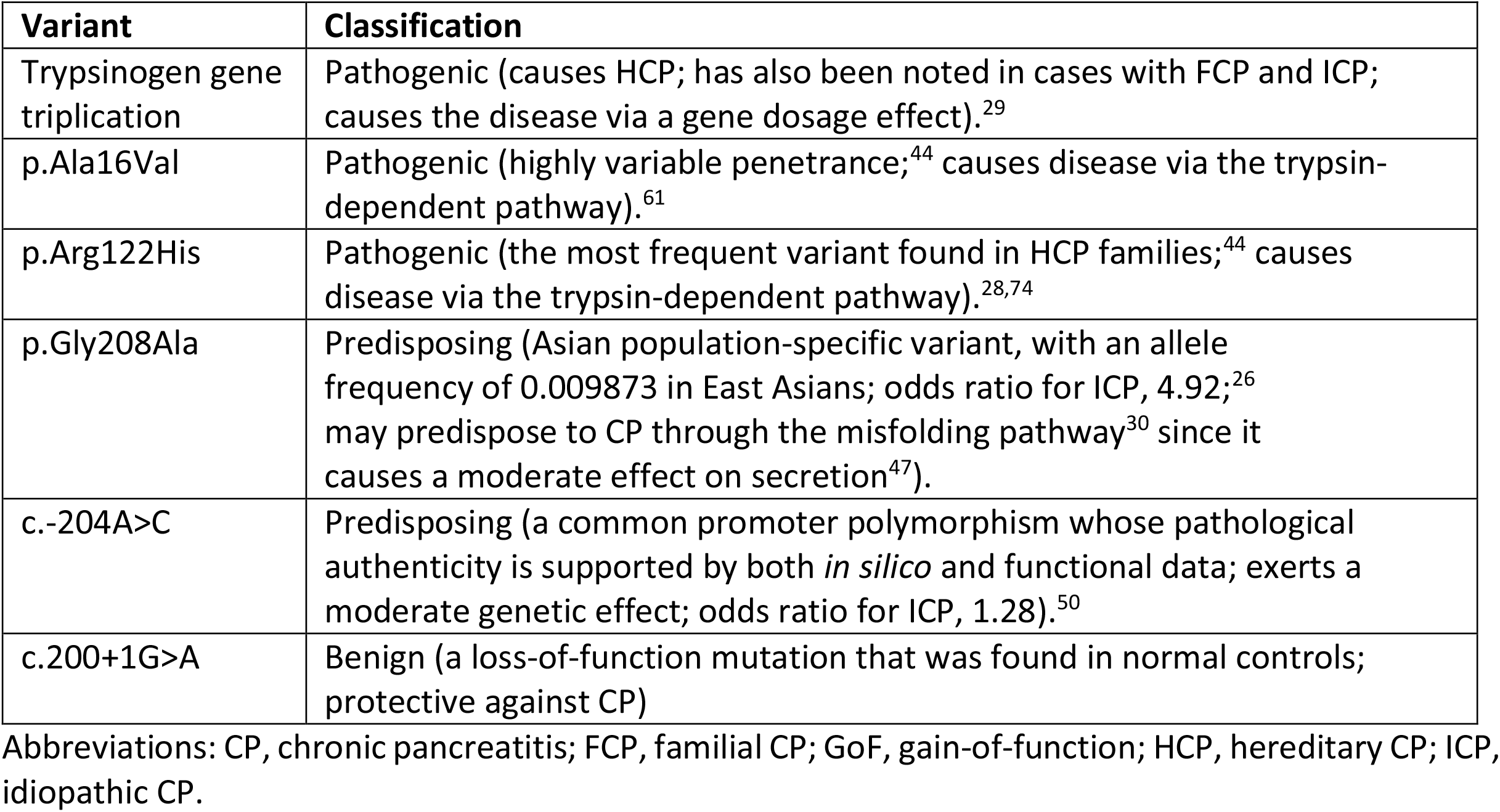
Illustrative examples of additions to the main classification categories in the context of *PRSS1* variants.

In the context of *SPINK1*, there would be three notable reclassifications. First, the abovementioned c.194+2T>C should be reclassified from “pathogenic” to “predisposing”. Second, the extensively studied p.Asn34Ser variant should be reclassified from “likely benign” to “benign’.^51-53^ Third, the functional enhancer variant, c.-4141G>T, which is in extensive linkage disequilibrium with p.Asn34Ser,^51,53^ should be reclassified from “likely pathogenic” to “predisposing” owing to its hspAF of ∼0.01975 (South Asia). Additionally, a very rare *SPINK1* variant, p.Arg65Gln, which was shown to cause a ∼50% functional loss,^54,55^ would be also reclassified from “pathogenic” to “predisposing” based upon the functional threshold (functional loss of >10 -<95%).

### Further additions to the classification framework

As mentioned above, it is desirable to provide necessary information about the pathologically relevant variant under study in parentheses immediately after the variant’s principal classification. The main reason is that, for any disease gene, there is often a large number of variants classified as either “pathogenic” or “predisposing”. We provide illustrative examples in the context of *PRSS1* variants in Table 6.

## Discussion

Using CP as a disease model and focusing on the four firmly established CP genes, we propose a general variant classification framework that both complements and extends the five ACMG-recommended categories (Figure 2). To this end, the first step taken was to classify the pathologically relevant variants in the different genes into three functional categories, GoF, LoF and GoP. This allowed us to appropriately perform several cross-gene and cross-variant comparisons, which then enabled us to assign the different genes into two distinct categories in terms of causality; causative genes refer to those in which a severe variant can cause CP on its own whilst predisposing genes refer to those in which even a highly deleterious variant cannot itself cause CP. This dichotomy is pivotal because it paves the way to extend or adapt the ACMG-AMP guidelines. Herein, we would like to emphasize that, in common with many term definitions, our currently defined “CP-causing genes” and “CP-predisposing genes” are context-dependent. Thus, we did not consider *CFTR* or *CTRC* as CP-causing genes even if compound heterozygous variants in both of them might cause CP.

Another key feature of our proposed conceptual framework was the adoption of two thresholds (allele frequency and functional) to separate pathogenic variants from predisposing variants in the disease-causing genes, thereby addressing the basic questions raised by Wright et al.^3^ We readily concede that the threshold values we settled upon, particularly the functional ones, may have to be adjusted once more data become available. In this regard, we would like to emphasize that the allele frequency and functional thresholds (and dichotomization of genes into disease causing and predisposing) would need to be established on a gene-by-gene basis and would require close collaboration between researchers and clinicians with expertise in the diseases/genes in question.

The salient point was that it was found to be unnecessary to make more than minimal changes to the five ACMG-AMP variant classification categories. As such, all the principles and rules established by ACMG-AMP may be readily used or adapted for variant classification using our proposed framework.

In summary, we propose a general variant classification framework that successfully addresses some basic questions about variant interpretation in human medical genetics. The maximal compliance of our proposed five-category and seven-category schemes (for disease-predisposing and causing genes, respectively) with the ACMG-AMP guidelines renders them readily applicable for variant classification in other well-established disease genes.

## Methods

### Variants in the four CP genes considered here

For reported variants in the *PRSS1, SPINK1* and *CTRC* genes, the reader should refer to the Genetic Risk Factors in Chronic Pancreatitis Database.^44^ Variants in the *CFTR* genes were sought in PubMed using a keyword search (i.e., *CFTR* plus pancreatitis plus variant or *CFTR* plus pancreatitis plus mutation; the latest search was performed on 12 April 2022). Data from some original reports were reinterpreted in accordance with the disease subtype definitions outlined below.

### Disease subtype definitions

CP empirically shown to have a genetic contribution may be classified into four subtypes, namely hereditary CP (HCP), familial CP (FCP), idiopathic CP (ICP) and alcoholic CP (ACP). The first three subtypes were defined in accordance with our previous practice. Specifically, HCP is defined in terms of having three or more affected family members spanning at least two generations whereas FCP is indicated by a positive family history but without satisfying the strict diagnostic criteria for HCP; ICP is specified when neither a positive family history of pancreatitis nor any obvious external causative risk factors (e.g., excessive alcohol consumption, infection, trauma and drug use) have been reported.^16,17,29^ ACP was defined in accordance with the original publications, in which it was usually attributed to an alcohol intake of ≥80 g/d for a male and 60 g/d for a female for at least two years. ‘Non-alcoholic CP’, a term used in some publications, is generally regarded as being equivalent to ICP, and this has been our previous practice.^41^ Finally, it should be emphasized that ICP was defined in terms of the absence of any identifiable etiology prior to genetic analysis.

### Variant allele frequency in normal populations

The Genome Aggregation Database (gnomAD; https://gnomad.broadinstitute.org/) was used (gnomAD v2.1.1 for missense and small insertion or deletion variants; gnomAD SVs v2.1 for copy number variants) as the source of variant population allele frequency data.^56^ Global population allele frequency (gpAF) refers to that calculated from all gnomAD populations. If a clinically identified variant is present in gnomAD, its highest subpopulation allele frequency (hspAF) was further evaluated. It should be noted that for variants in the *PRSS1* gene, we determined whether they could represent sequence artifacts as in the case of p.Ala16Val.^57^

### Observed/expected (o/e) scores of predicted loss-of-function (pLoF) variants

The pLoF o/e scores for the four CP genes were obtained from gnomAD v2.1.1.^56^

### Protein tissue expression data

Protein tissue expression data were in accordance with the Human Protein Atlas (https://www.proteinatlas.org).

## Data Availability

All data produced in the present work are contained in the manuscript

## Data availability

All supporting data are available within the article or its supplementary materials.

## Acknowledgments

This work was performed within the framework of the Sino-French GREPAN (Genetic REsearch on PANcreatitis) study and was supported by the Institut National de la Santé et de la Recherche Médicale (INSERM), the Association des Pancréatites Chroniques Héréditaires and the Association Gaétan Saleün, France and the National Natural Science Foundation of China (no. 82120108006 [Z.L.]). N.P. received a one-year visiting PhD student scholarship from the China Scholarship Council, the Ministry of Education of China (no. 202006190267). The funding sources did not play any role in the study design, collection, analysis or interpretation of the data or in the writing of the report.

## Author contributions

E.M. and W-B.Z. contributed to the study design, data collation and analysis, and assisted in writing the paper. E.G., D.N.C., G.L.G., Y.F., N.P. and V.R. analyzed the data and critically revised the manuscript with important intellectual input. C.F. and Z.L. contributed to the concept of the study, analyzed the data and critically revised the manuscript. J.M.C. conceived and coordinated the study, performed data collation and analysis, and drafted the manuscript. All authors approved the final draft submitted.

